# An AI-Supported Methodology for Identifying Attachment Styles

**DOI:** 10.1101/2025.08.30.25334439

**Authors:** Yair Neuman, Keren Hazdai

## Abstract

Identifying attachment styles is important for clinical psychologists interested in better understanding their patients. Traditional methods for identifying attachment styles rely on clinical interviews or questionnaires, which are time-consuming and subject to observer bias. Large Language Models offer potential for automated, objective attachment style analysis. The aim of the paper was to develop and validate an AI-supported methodology for identifying attachment styles using the four-category model. To validate the methodology, we analyzed the narratives of a unique group of subjects who experienced an acute psychological crisis. A LLM was used to analyze each stage using a structured prompt. We hypothesized that if the methodology validly identifies attachment styles, the fearful style would be the most prevalent in the descriptions of the subjects’ insecure childhood and crises. The methodology successfully identified the theoretically predicted attachment styles. Chi-square analyses confirmed significant deviations from the null hypothesis across all stages with a strong effect size. In sum, the AI-supported attachment analysis offers an objective, efficient alternative to traditional methods while maintaining theoretical validity. The methodology demonstrates potential clinical utility for rapid assessment of attachment dynamics, though further validation across diverse populations is warranted.

## Introduction

The importance of identifying attachment styles for understanding clinical processes is well-established (e.g., [1–5]) and can be extended to the dynamic analysis of these styles, specifically for understanding an emerging crisis (e.g., [6–8]). Attachment styles are often identified either qualitatively, through a clinical interview, or through questionnaires. However, advancements in AI, specifically in the development of Large Language Models (LLMs), may introduce a new, quick, and valid approach for automatically identifying attachment style through the analysis of naturalistic data. Such an approach is grounded in the success of LLMs to validly analyze human interactions, specifically for a psychological analysis (e.g., [9]) and concerning the sensitive contextual analysis required from such an interpretation [10]. This short paper aims to introduce and provide preliminary validation of an AI-supported methodology for identifying attachment styles. The validation is done through a unique dataset of interviews with state witnesses.

State witnesses are senior criminals who have decided to switch sides and testify against their fellow members in the crime organization. To validate the proposed methodology, we analyzed in-depth interviews conducted with 13 state witnesses. Based on a previous qualitative psychodynamic analysis of the witnesses’ narratives [11], from childhood up to their decision to cross the lines, we hypothesized that a fearful attachment style characterizes the childhood of these criminals. The basic sense of insecurity underlying this style is addressed by joining the criminal world and gaining a secure base (i.e., a secure attachment style), reaching a peak at the height of their criminal career. However, repeating crises draw them back into the fearful attachment style, reaching its apex in an event (i.e., the triggering event) that has led them to betray their organization, while giving up their association and status and risking their life. As the witnesses experience a crisis concerning their decision to testify against their crime organization, we expected a valid AI methodology to identify the attachment types corresponding with different chronological stages of the witnesses’ narratives.

## Method

### Participants

This study is based on semi-structured, in-depth interviews with 13 state witnesses. The sample represented the entire population of state witnesses during the relevant timeframe. These subjects were selected in coordination with law enforcement authorities and the Witness Protection Authority, and the recruitment process received formal approval from the Ministry of National Security, which also supervised the interviews. Written and signed informed consent was obtained from each participant. Institutional Review Board (IRB) approval was obtained from the Government Office responsible for witness protection (ID: 08384917). The interview protocols were approved by the Bar-Illan University ethics committee (Date: 1.2.2018, No document ID). The second author, a Ph.D. student at Bar-Illan Univ., collected the data then.

### Procedure

The research design was qualitative and longitudinal. Two expert clinicians interviewed the participants individually and asked them to recount their life histories from early childhood to their involvement in their crime organization. The interviews then focused on the transitional period in which participants shifted from being members of a criminal organization to becoming state witnesses. For those who had already provided testimony, additional focus was placed on the post-transition period. Interviews ranged from approximately 2.5 to 6 hours, incorporating one or more breaks depending on the interviewee’s needs. All interviews were audio-recorded and fully transcribed verbatim for subsequent analysis.

### Data Analysis

A narrative-based analysis approach was employed to segment each interview into chronological stages. A distinct chronological pattern emerged during the initial reading of the transcripts, reflecting a consistent sequence of life events across participants. The convergence in narrative structure and interpretation allowed the data to be segmented into ten key temporal units, each representing a core phase in participants’ stories.

### Chronological Stages Identified

The interviews were segmented into the following ten developmental phases:

#### 1. Childhood (Birth–Age 12)

Descriptions of early attachment styles and relational experiences were central. Participants frequently linked adverse childhood events to the development of personality psychopathology and a later predisposition toward criminal behavior.

#### 2. Early Adolescence (Ages 12–14)

This stage marked the beginning of identity differentiation and social deviation, including initial associations with marginal peer groups and the development of a nascent sense of autonomy and agency.

#### 3. Adolescence (Ages 14–18)

A critical period in which a criminal identity became consolidated. Participants described escalating involvement in illegal activities and deepening identification with delinquent peer networks.

#### 4. Peak Criminal Adulthood

Representing the apex of criminal careers—spanning years or even decades— this stage involved robust criminal identities, reinforced by success stories and emotionally fulfilling bonds within the criminal milieu.

#### 5. First Crisis

A significant turning point marked by internal or interpersonal conflicts that disrupted the integrity of the criminal identity. This stage often introduced the first elements of doubt or disillusionment.

#### 6. Resolution of First Crisis

A phase of re-alignment, during which participants managed to reintegrate into the criminal environment through adaptive strategies that restored a sense of stability and control.

#### 7. Second Crisis

A more profound, often violent rupture in the relationship with the criminal organization. This stage was frequently described in terms of escalating threats, violence, or existential danger.

#### 8. Resolution of Second Crisis

Characterized by a tense and partial disengagement from key roles within the criminal organization. Participants described this period as involving compromise, emotional exhaustion, and partial withdrawal.

#### 9. Context for the Trigger

This phase encompassed symbolic or emotionally charged events that provided the psychological context for subsequent defection. These events often laid the groundwork for reconsidering allegiance to the criminal organization.

#### 10. Triggering Event

The final, decisive moment—often seemingly trivial—initiated a profound internal shift. This event was experienced as a moment of emotional clarity and insight, ultimately catalysing the decision to cooperate with law enforcement and become a state witness.

### Identifying attachment styles

We used the four-type category model to identify attachment styles expressed in each chronological stage [12]. We separately analyzed each chronological stage using ChatGPT as the Large Language Model. To design the prompt, we used another LLM (i.e., Claude) and asked the model to read [12]’s paper and to design a prompt for identifying attachment styles. It is important to emphasize that, following the state-of-the-art in the field (e.g., [13]), we designed the prompt by integrating our experts’ knowledge with the proposal of the AI. The prompt used for the analysis is:

***

### Attachment Style Analysis Prompt

(Based on Bartholomew & Horowitz, 1991) Please read and analyze the text below through the lens of Bartholomew’s four-category attachment theory. This model identifies four attachment styles based on two orthogonal dimensions: Model of Self (positive/negative) and Model of Others (positive/negative).

### Analysis Instructions

#### Step 1: Identify Relevant Content

Look for specific descriptions of the SELF and OTHERS only. Focus on: Self-descriptions - Direct or implicit characterizations of the narrator:

- Personal qualities, traits, or characteristics
- Self-worth, lovability, or value as a person
- Capabilities, competencies, or inadequacies
- Identity or self-concept statements

OTHERS descriptions - Direct or implicit characterizations of other people (especially caregivers):

- Personal qualities, traits, or characteristics of others
- Trustworthiness, reliability, or supportiveness of others
- How others treat or regard the narrator
- Others’ emotional availability or responsiveness

EXCLUDE:

- Situational descriptions (events, circumstances, processes)
- Behavioral descriptions without character implications
- Factual information about locations, timing, or logistics
- General relationship dynamics without specific person characterizations

#### Step 2: Categorize and Score

For each identified specific description of SELF or OTHERS:

1. Categorize as either Self-Model or Other-Model
2. Quote the exact phrase that describes the person/people
3. Determine valence:
  ∘ Positive Self-Model (+): Self described as worthy, capable, lovable, confident, valuable
  ∘ Negative Self-Model (-): Self described as unworthy, incapable, unlovable, damaged, flawed
  ∘ Positive Other-Model (+): Others described as trustworthy, caring, supportive, reliable, available
  ∘ Negative Other-Model (-): Others described as harmful, rejecting, unreliable, uncaring, threatening

4. Assign valence score: −1.0 (highly negative) to +1.0 (highly positive) Important: Only score actual descriptions of what people ARE like, not what they DO or what happens to them.

#### Step 3: Calculate Averages

- Average Self-Model score
- Average Other-Model score

Step 4: Determine Attachment Style

Based on the combination of positive (+) or negative (-) averages:

| Self-Model | Other-Model | Attachment Style | Characteristics |
| --- | --- | --- | --- |
| Positive | Positive | SECURE | Comfortable with intimacy and autonomy |
| Negative | Positive | PREOCCUPIED | Seeks acceptance from others; relationship-focused |
| Positive | Negative | DISMISSING | Values independence; avoids close relationships |
| Negative | Negative | FEARFUL | Desires closeness but fears rejection |

#### Step 5: Supporting Evidence

Identify specific behavioral or emotional patterns that support the classification:

- Secure: Coherent relationship descriptions, balanced autonomy/intimacy
- Preoccupied: Excessive disclosure, emotional volatility, idealization of others
- Dismissing: Emotional restriction, emphasis on self-reliance, devaluation of relationships
- Fearful-Avoidant: Social anxiety, approach-avoidance conflicts, fear of rejection

#### Output Format

Self-Model Descriptions:

- [Quote exact phrase] - Score: [X.XX] - [Brief rationale focusing on character description]
- Average Self-Model Score: [X.XX]

Other-Model Descriptions:

- [Quote exact phrase] - Score: [X.XX] - [Brief rationale focusing on character description]
- Average Other-Model Score: [X.XX]

Predicted Attachment Style: [STYLE NAME]

Supporting Evidence:

- [Key behavioral/emotional patterns observed]
- [Specific quotes or examples that exemplify the style]

Confidence Level: [High/Medium/Low] with brief explanation

***

We applied this prompt to the analysis of the interviews and for each chronological stage, generated its dominant attachment style. For example, we analyzed the interview with “Eric” (a pseudonym), a state witness with clear psychopathic traits, who was a leading figure in the criminal world. In his childhood description, he says:

> “I grew up with parents who were both former military personnel. Until the age of XXX, we lived in XXX, and then moved to XXX (a military base), followed by a year in XXX, and afterward to XXX. In XXX, my parents began the process of divorce and continued it in XXX. The divorce dragged on for seven years. We were exposed to violence, especially me, mostly from my parents.”

The model scores the description of self and others as follows:

SELF = −0.90

OTHERS = −0.70

Moreover, the attachment style was identified as FEARFUL-AVOIDANT.

### Hypotheses

We hypothesized that if the methodology validly identifies attachment styles, the FEARFUL style would be the most prevalent in childhood, crises, and triggering events. In contrast, the SECURE attachment style would be the most prevalent in the Peak Criminal Adulthood stage. The justification for these hypotheses has been previously explained.

## Results

Using the above-mentioned prompt, we analyze the chronological time units (i.e., the stages). Table 1 presents the distribution of attachment style by stage: childhood, peak, crisis 1, crisis 2, and the triggering event. The table presents the results in terms of rounded percentages:

**Table 1.** The distribution of attachment styles by stage.

| Stage | FEA. | DIS. | SEC. | PRE. |
| --- | --- | --- | --- | --- |
| **CHILD** | 76 | 8 | 8 | 8 |
| **PEAK** | 0 | 8 | 92 | 0 |
| **CRISIS 1** | 62 | 23 | 0 | 15 |
| **CRISIS 2** | 85 | 8 | 0 | 7 |
| **TRIGGERING** | 100 | 0 | 0 | 0 |

For each stage, the observed distribution of attachment styles differs significantly from the one expected under the null hypothesis. In childhood, the dominant style is fearful (χ^2^(3) = 18.70, p <.001), in the peak of the criminal career, the dominant style is secure (χ^2^(3) = 31.62, p <.001), in the crises and the triggering events that led to the betrayal, the dominant style is fearful again (χ^2^(3) = 10.69, p < 0.05, χ^2^(3) = 24.85, p < 0.001, andχ^2^(3) = 39.00, p < 0.001 respectively).

Cramér’s V was calculated to complement the chi-square tests by measuring the effect size. The effect sizes were large for all conditions: childhood (V =.69), peak (V =.90), crisis 1 (V =.52), crisis 2 (V =.80), and triggering (V = 1.00), indicating strong associations between attachment style and phase.

These results support our hypotheses and validate the methodology. Moreover, analyzing the dynamics of attachment styles, we measured the Shannon entropy associated with each stage; the lower the entropy, the more concentrated the distribution is around a dominant attachment style. Figure 1 presents the results:

**Figure 1.**
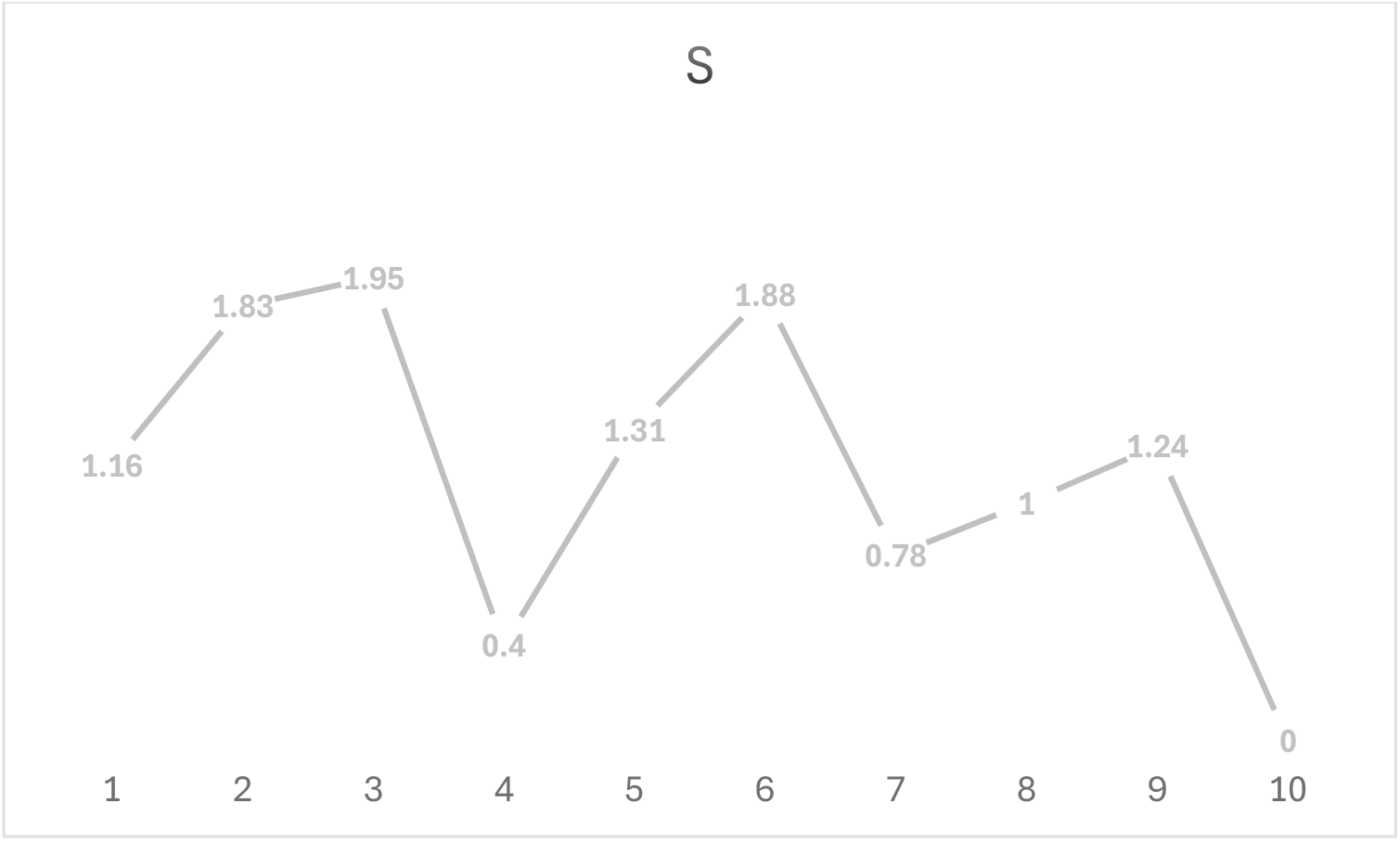
The entropy of attachment styles as a function of chronological stage.

We can see that the entropy score significantly changes as a function of chronological stage. At the peak of their criminal career, the state witnesses describe a reality dominated mainly by a secure attachment style. However, describing the triggering experience that led them to cross the lines, there is an absolute dominance of the fearful style.

## Discussion

Psychiatrists and clinical psychologists interested in attachment may extend their tool kit using the methodology proposed in this paper. The methodology has two main benefits. First, it is objective because human bias is removed from the analysis. Second, it is time and energetically efficient. Moreover, we used a specific attachment model to analyze the interviews. However, LLMs such as ChatGPT or Claude may be instructed to read about different attachment models and use them to design different prompts adhering to the different models.

The benefits of using the proposed methodology do not outweigh the possible difficulties of using LLMs. However, previous concerns regarding hallucinations can be sufficiently addressed through the appropriate design of prompts, the model’s recursive analysis of its own product (e.g., [13]), and external validation. In sum, the proposed methodology may enrich the toolkit of psychiatrists and clinical psychologists sensitive to the transformative power of innovative technologies while acknowledging the power of theories and clinical knowledge to recursively inform the use of these technologies.

Our study validated the methodology using a specific sample. However, the clinical implications of this AI-supported attachment analysis methodology extend beyond its demonstrated validity in a specific forensic population. The methodology’s ability to detect attachment style transitions, as evidenced by the shift from fearful attachment in childhood to secure attachment during peak criminal careers, suggests potential utility in monitoring therapeutic progress and identifying periods of vulnerability. Recording a session with a client and analyzing it online may give the psychotherapist a powerful tool for monitoring and regulating the process.

The predominance of fearful attachment during crisis periods and the triggering event (85% and 100% respectively) may suggest that individuals with histories of fearful attachment may be particularly susceptible to withdrawing under stress, a finding with direct relevance to crisis intervention and relapse prevention strategies. The methodology’s power to identify these vulnerable periods through narrative analysis could inform risk stratification protocols, particularly in forensic psychiatric settings where understanding attachment dynamics is crucial for both treatment planning and risk assessment.

From a broader diagnostic perspective, the proposed AI-based methodology offers potential advantages in populations where traditional attachment assessments may be difficult or impossible to apply. Patients with personality disorders, particularly those with antisocial or borderline presentations, often demonstrate limited insight or may consciously distort self-reporting on standardized measures. The narrative-based approach, analyzed through LLM, may address some limitations by identifying attachment patterns embedded within naturalistic discourse, potentially revealing unconscious or defended attachment representations. However, beyond the previously discussed qualifications, several additional limitations warrant consideration before clinical implementation. The methodology requires validation across diverse populations, as the current sample’s unique characteristics may not generalize to other populations. Moreover, relying on narrative assumes adequate verbal expression capabilities, potentially limiting applicability in populations with thought disorders, cognitive impairment, or cultural/linguistic barriers. Nevertheless, researchers and practitioners who would run the prompt on their data may immediately sense the face validity of the proposed methodology. Trained on a vast corpus of human language and context-sensitive, LLMs produce excellent results when accurately prompted to analyze human expression of thought processes expressed through language.

## Data Availability

Interviews are available upon request

## Declaration of ethics

All relevant ethical guidelines have been followed, all necessary IRB and/or ethics committee approvals have been obtained, all necessary patient/participant consent has been obtained, and the appropriate institutional forms have been archived.

## References

1. Bowlby J. A secure base. London: Routledge, 2012.

2. Bowlby J, Bowlby R. The making and breaking of affectional bonds. London: Routledge, 2012.

3. Holmes J. The search for the secure base: Attachment theory and psychotherapy. London: Routledge, 2014.

4. Holmes J. Exploring in security: Towards an attachment-informed psychoanalytic psychotherapy. London: Routledge; 2009.

5. Holmes J. Attachment theory in clinical practice: A personal account. Br J Psychother. 2015;31(2):208–28.

6. Chopik WJ, Edelstein RS, Grimm KJ. Longitudinal changes in attachment orientation over a 59-year period. J Pers Soc Psychol. 2019;116(4):598–615.

7. Fraley RC, Roisman GI. The development of adult attachment styles: Four lessons. Curr Opin Psychol. 2019;25:26–30.

8. Fraley RC, Gillath O, Deboeck PR. Do life events lead to enduring changes in adult attachment styles? A naturalistic longitudinal investigation. J Pers Soc Psychol. 2021;120(6):1567–82.

9. Neuman Y, Cohen Y. A data set of synthetic utterances for computational personality analysis. Sci Data. 2024;11(1):623.

10. Neuman Y. AI for understanding context. New York: Springer; 2024. (SpringerBriefs in Computer Science).

11. Authors. Latent vulnerability as a predisposition for betrayal. Under review.

12. Bartholomew K, Horowitz LM. Attachment styles among young adults: A test of a four-category model. J Pers Soc Psychol. 1991;61(2):226–44.

13. Neuman Y. AI for understanding human conversations. Boca Raton, FL: CRC Press/Routledge; 2025. (AI for Everything).

